# The relationship between autoantibodies targeting GPCRs and the renin-angiotensin system associates with COVID-19 severity

**DOI:** 10.1101/2021.08.24.21262385

**Authors:** Otavio Cabral-Marques, Gilad Halpert, Lena F. Schimke, Yuri Ostrinski, Israel Zyskind, Miriam T. Lattin, Florian Tran, Stefan Schreiber, Alexandre H.C. Marques, Igor Salerno Filgueiras, Desirée Rodrigues Plaça, Gabriela Crispim Baiocchi, Paula Paccielli Freire, Dennyson Leandro M. Fonseca, Jens Y. Humrich, Tanja Lange, Antje Müller, Lasse M. Giil, Hanna Graßhoff, Anja Schumann, Alexander Maximilian Hackel, Juliane Junker, Carlotta Meyer, Hans D. Ochs, Yael Bublil Lavi, Kai Schulze-Forster, Jonathan I. Silvergerg, Howard Amital, Jason Zimmerman, Harry Heidecke, Avi Z Rosenberg, Gabriela Riemekasten, Yehuda Shoenfeld

**Author notes:** **Corresponding author: Yehuda Shoenfeld**, MD, FRCP, MaACR, Zabludowicz Center for Autoimmune Diseases, Sheba Medical Center, affiliated with the Sackler, Faculty of Medicine, Tel-Aviv University, Tel-Hashomer 5265601, Israel, **Otavio Cabral-Marques**, MSc, PhD, Department of Immunology, Institute of Biomedical Sciences - University of São Paulo, Lineu Prestes Avenue, 1730, São Paulo, SP, 05508-900, Brazil, **Gabriela Riemekasten**, MD, Department of Rheumatology and Clinical Immunology, University of Lübeck, Lübeck, 23538, Germany, Klinik für Rheumatologie, Ratzeburger Allee 160 (Haus 40), 23562 Lübeck. Contributed equally (Otavio Cabral-Marques, Gilad Halpert, Lena F. Schimke, Gabriela Riemekasten, Yehuda Shoenfeld).

## Abstract

The coronavirus disease 2019 (COVID-19) can evolve to clinical manifestations resembling systemic autoimmune diseases, with the presence of autoantibodies that are still poorly characterized. To address this issue, we performed a cross-sectional study of 246 individuals to determine whether autoantibodies targeting G protein-coupled receptors (GPCRs) and renin-angiotensin system (RAS)-related molecules were associated with COVID-19-related clinical outcomes. Moderate and severe patients exhibited the highest autoantibody levels, relative to both healthy controls and patients with mild COVID-19 symptoms. Random Forest, a machine learning model, ranked anti-GPCR autoantibodies targeting downstream molecules in the RAS signaling pathway such as the angiotensin II type 1 and Mas receptor, and the chemokine receptor CXCR3 as the three strongest predictors of severe disease. Moreover, while the autoantibody network signatures were relatively conserved in patients with mild COVID-19 compared to healthy controls, they were disrupted in moderate and most perturbed in severe patients. Our data indicate that the relationship between autoantibodies targeting GPCRs and RAS-related molecules associates with the clinical severity of COVID-19, suggesting novel molecular pathways for therapeutic interventions.

## MAIN

Autoantibodies have been identified in patients with coronavirus disease 2019 (COVID-19), suggesting that the infection by severe acute respiratory syndrome virus 2 (SARS-CoV-2) can evolve to a systemic autoimmune disease^1–5^. For instance, high levels of antiphospholipid autoantibodies have been linked to severe respiratory disease by inducing neutrophil extracellular traps (NETs) and venous thrombosis^4,6–9^. High titers of neutralizing immunoglobulin G (IgG) autoantibodies against type I interferons (IFNs) have been reported in patients with life-threatening COVID-19^10^. Most recently, a wide range of autoantibodies in patients with COVID-19 have been characterized using rapid extracellular antigen profiling (REAP)^11^, a technology for comprehensive and high-throughput identification of autoantibodies recognizing 2,770 extracellular and secreted protein components of the exoproteome (extracellular protein epitopes)^12^. Wang, E. Y. *et al*^11^ showed that both healthy controls and COVID-19 patients have multiple autoantibodies against the exoproteome. While all patients with COVID-19 displayed reactivity against a larger number of proteins, those with severe disease had the highest reactive scores.

These results are in line with our previous report^13^ on autoantibodies targeting the largest superfamily of integral membrane proteins in humans^14^, i.e., the G protein-coupled receptors (GPCRs) suggesting that these autoantibodies are natural components of human biology that become dysregulated in autoimmune diseases. Likewise, recent studies have detected functional antibodies against GPCRs in the sera of patients with COVID-19 and indicate that they may be associated with disease severity^15–17^. However, these investigations were not systemic, focusing only on two types of anti-GPCR autoantibodies and did not investigate their relationship with the potential presence of autoantibodies targeting molecules of the immune and the renin-angiotensin systems (RAS), which play a central role in the development of severe COVID-19.

## RESULTS

### Autoantibodies against GPCRs and renin-angiotensin system (RAS)-related molecules

Here, we investigated the serum levels of autoantibodies targeting molecules belonging to the RAS (including GPCRs: MASR, AT1R, and AT2R as well as the entry receptor for the SARS-CoV-2, Angiotensin-converting enzyme II [ACE-II])^18–21^. Furthermore, we assessed the concentrations of autoantibodies against GPCRs involved in chemotaxis and inflammation (CXCR3^22,23^ and C5aR^24^), coagulation (PAR1^25^), and neuronal receptors (ADRA1A, ADRB1, and ADRB2, ACHRMs)^26–30^, which have been implicated in the development of COVID-19 disease (see **Supp. Table 1** for abbreviations of autoantibodies and their targets). In addition, we investigated autoantibodies targeting receptors facilitating the infectivity of SARS-CoV-2, and its entrance into host cells (neuropilin-Ab)^31^. Finally, we explored the potential presence of autoantibodies against STAB1 (STAB-1-Ab) as a potential new candidate involved in COVID-19 infectivity, which is a scavenger receptor still not investigated for any role in COVID-19. However, its multifunctionality during leukocyte trafficking, tissue homeostasis, and resolution of inflammation suggests it could be relevant for disease severity^32,33^. **Figure 1A** and **Figure 1B** represent how these autoantibody targets are interconnected by protein-protein interaction (PPI) or gene ontology (GO) relationships, respectively.

**Figure 1:**
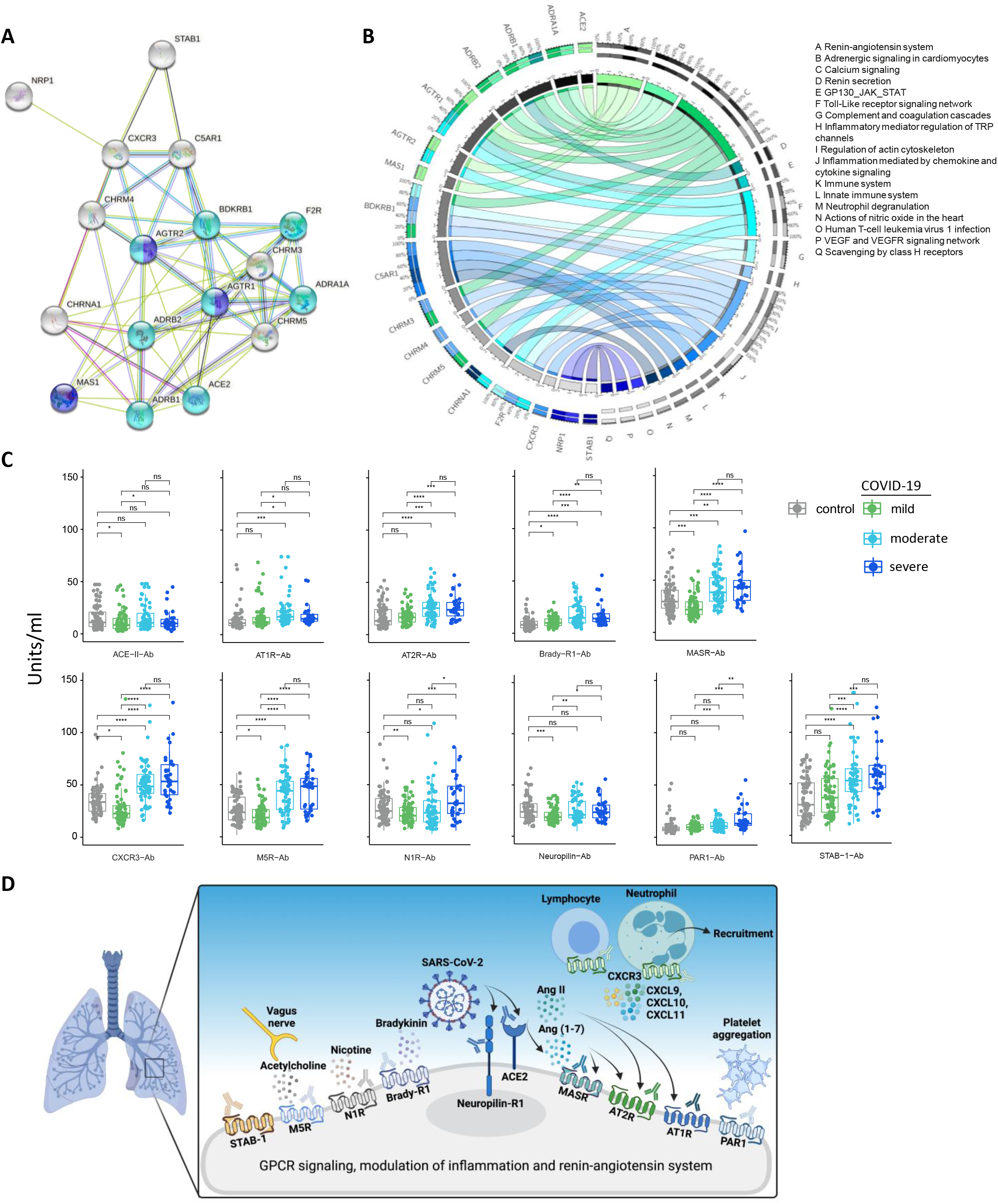
Autoantibodies against GPCRs and COVID-19-associated molecules are significantly different between healthy controls and COVID-19 patients. Interaction network of anti-GPCR-autoantibody targets. Purple molecules represent GPCRs involved in the angiotensin-activated signaling pathway and blue are those involved in regulation of blood pressure. Grey are molecules belonging to other gene ontology categories. Line color indicates the type of interaction evidence: dark-green represents gene neighborhood, light-green denotes text mining, black indicates co-expression, blue shows gene co-occurrence, purple evidences protein homology, and pink marks experimental determined. (**B**) Circos plot illustrates the functional relationship between the antibody targets and their biological functions as indicated by gene ontology (GO) enriched terms (denoted by letters and legends).Complete list of associations are described in **Supp. Table 1**. Thickness of rectangles in each small outer circles is proportional to their involvement of autoantibody targets and in multiple pathways. The inner circle represents genes and datasets with more connections to each other. The colors, numbers and percentage on the outer circles denote how pleiotropic and the respective each gene/pathway association. (**C**) Box plots of 11 antibodies with significantly different expression levels (illustrated in **D** with functional associations of their targets) in at least one group of COVID-19 patients (mild, moderate or severe group) compared to healthy controls. Significant differences between groups are indicated by asterisks (* p ≤ 0.05, ** p ≤ 0.01 and *** p ≤ 0.001).

We found significantly higher levels of autoantibodies directed against eleven receptors (**Figure 1C**), which are involved in the modulation of inflammation and the RAS (**Figure 1D**) suggesting an association between autoantibodies against GPCRs and RAS-related molecules with COVID-19 severity. In contrast, both controls and all COVID-19 disease groups were found to have a number of other autoantibodies at similar levels, most of them targeting neuronal receptors (ADRA1R, ADRB1, ADRB2, CHRM3, and CHRM4)^34–36^, but also against the receptor for complement C5a (C5aR), a potent anaphylatoxin chemotactic receptor^37^, suggesting that severe COVID-19 is specifically associated with autoantibodies towards certain groups of GPCRs (**Supp. Fig 1**).

### Autoantibody stratification of COVID-19 severity using multivariate analyses

Next, we carried out principal component analysis (PCA) using a spectral decomposition approach^38,39^, to examine the correlations between variables (autoantibodies) and observations (individuals) while stratifying groups based on the autoantibody levels. This approach indicated that autoantibodies stratify COVID-19 patients according to disease severity (mild, moderate, and severe patients) (**Figure 2A and 2C**). While healthy controls and patients with mild COVID-19 present a closer autoantibody pattern, moderate and severe COVID-19 patients clustered together. In this context, autoantibodies such as ACE-II-Ab, AT2R-Ab, Brady-R1, CXCR3-Ab, MASR-Ab, M5R, neuropilin-Ab, PAR1-Ab, STAB-1Ab appeared to play a major role in stratifying COVID-19 by disease burden (**Figure 2B-2D**). Altogether, these results indicate that the association between autoantibodies against GPCRs and COVID-19-related molecules can be used as biomarkers for COVID-19 burden.

**Figure 2:**
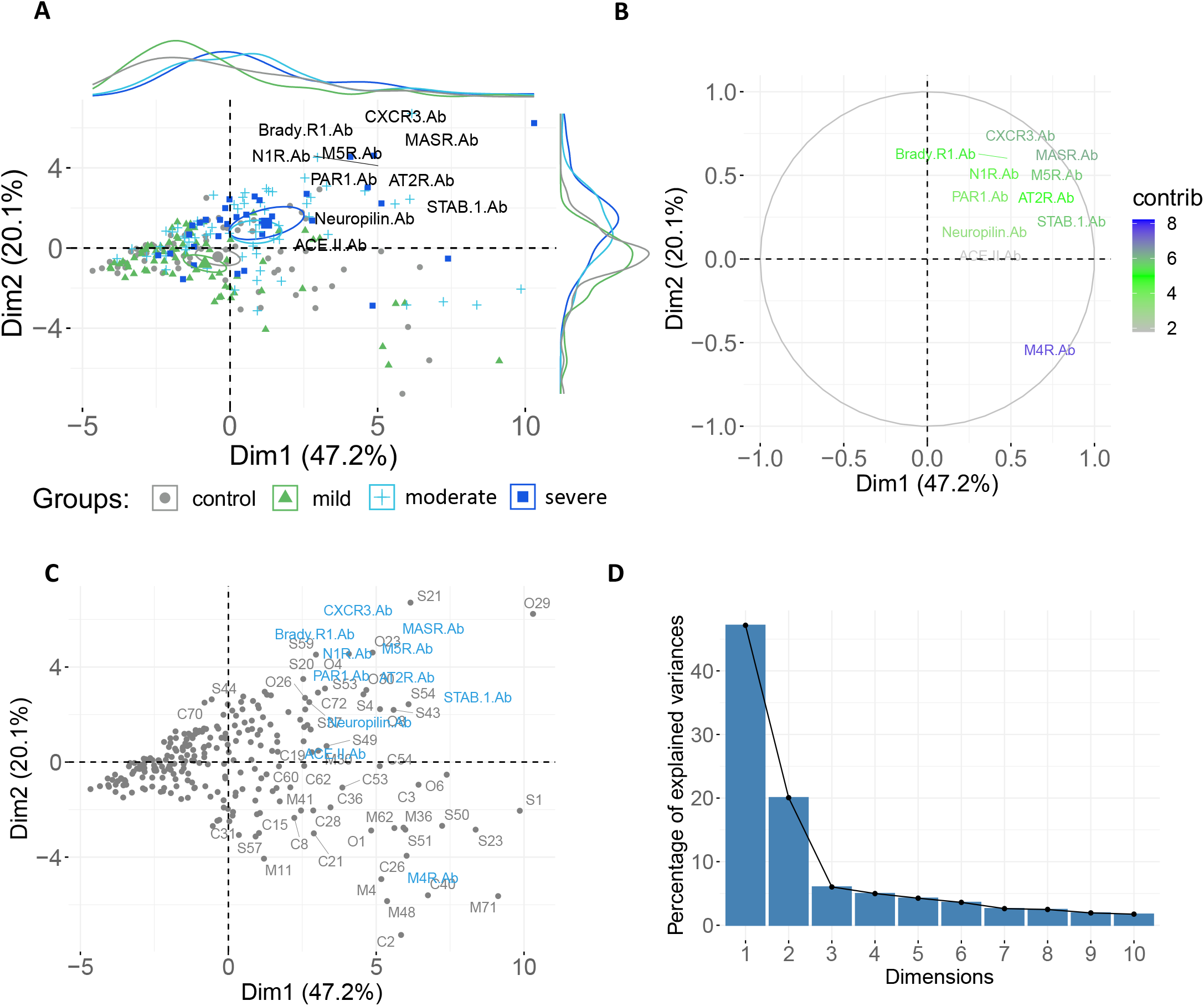
Autoantibodies stratify COVID-19 patients by disease severity. **(A)** Principal Component Analysis (PCA) with spectral decomposition based on 17 different anti-GPCR-autoantibodies shows the stratification of severe and oxygen-dependent COVID-19 patients from mild COVID-19 patients and healthy controls. Variables with positive correlation are pointing to the same side of the plot, contrasting with negative correlated variables, which point to opposite sides. Only highly contributing to the stratification of severe and oxygen-dependent patients from mild COVID-19 and healthy controls are shown. Confidence ellipses are shown for each group/category. **(B)** Graphs of variables (antibodies) obtained by PCA analysis of COVID-19 mild, moderate and severe group and healthy controls, indicating autoantibodies highly associated with moderate and severe COVID-19 **(C)** Biplot of individuals and variables of same groups as in **A**. Individuals with a similar profile are grouped together. **(D)** Scree plot showing the percentage of variances explained by each principal components (PCs). The x-axis shows the number of PCs and the y-axis the percentage of explained variances.

### Machine learning classification of COVID-19 patients based on autoantibodies

To further explore the potential of autoantibodies to predict COVID-19 outcomes, we performed Random Forest modelling, which is a machine learning approach that establishes outcomes based on predictions of decision trees^40^. The receiver operating characteristics (ROC) curve indicated a high false-positive rate for the classification of severe patients with the stable curve showing the highest error rate (out-of-bag or OOB) for this group (**Figure 3A** and **3B**). I.e., in accordance with the PCA analysis, Random Forest classification of COVID-19 groups showed a higher error rate (low accuracy) when distinguishing moderate patients from those with severe COVID-19.

**Figure 3:**
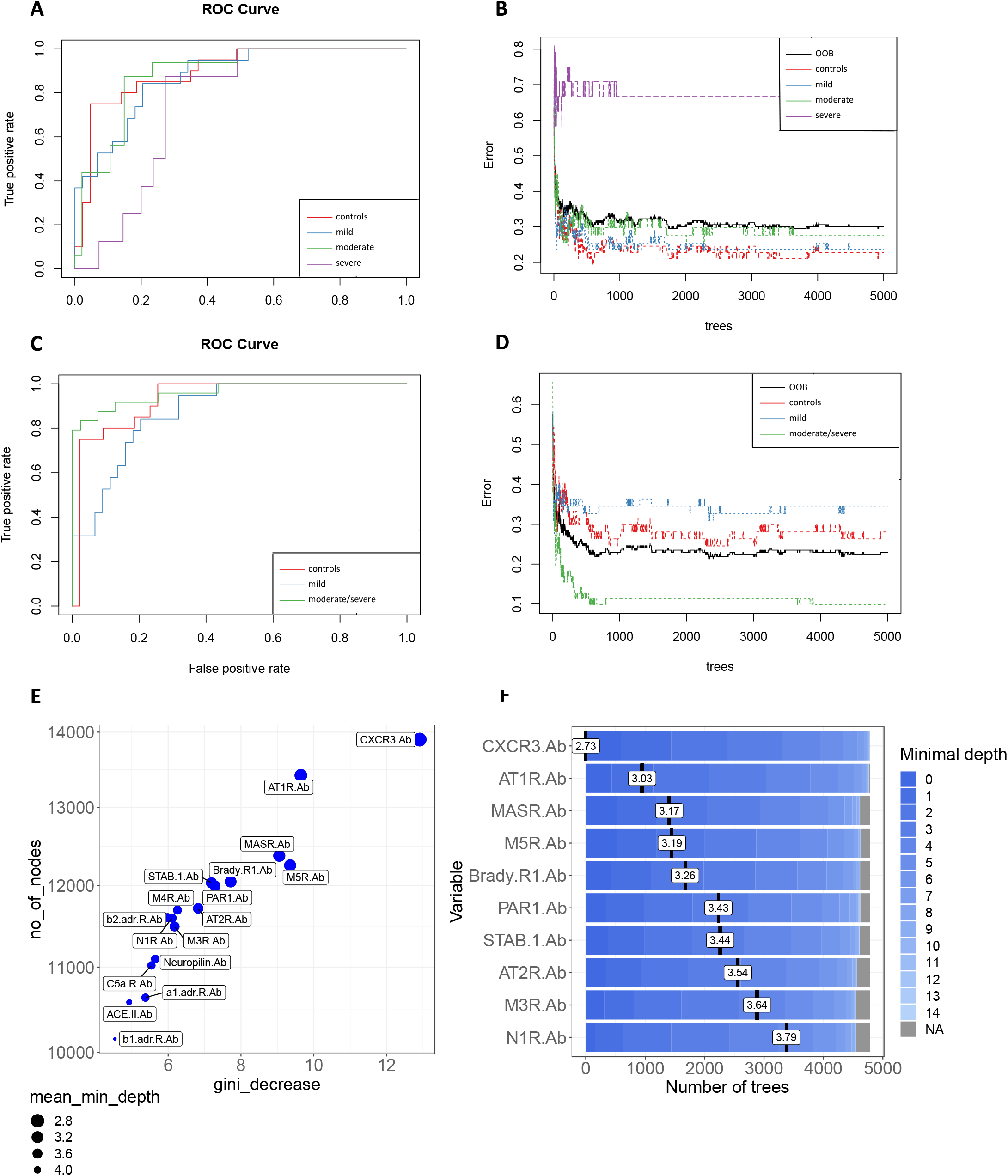
Ranking autoantibodies as predictors of COVID-19 severity. (**A**) Receiver operating characteristics (ROC) curve of 17 antibodies from COVID-19 mild, moderate, and severe patients compared to healthy controls. (**B**) Stable curve showing number of trees and out-of-bag (OOB) error rate of 30,05%.(**C**) ROC curve of the same antibodies as in (A) from mild COVID-19 and moderate/severe COVID-19 patients compared to healthy controls with an area under the curve (AUC) of 93,1% (for controls), 87,7% (for mild) and 96,2% (for moderate/severe), respectively. (**D**) Stable curve showing number of trees and OOB error rate of 22,95%. **(E)** Variable importance scores plot based on gini decrease and number(no) of_nodes for each variable showing which variable (antibody) presents a higher score to predict COVID-19 severity.

Thus, we assigned moderate and severe COVID-19 patients to the same group to identify the most relevant autoantibody predictors of COVID-19 burden. Using this approach, the merged moderate/severe patient group showed the lowest error rate compared to healthy controls and mild COVID-19 patients. This model resulted in an OOB error rate of 22,95% for all groups and an area under the ROC curve of 0.93, 0.87, and 0.96 for healthy controls, COVID-19 mild, and COVID-19 moderate/severe groups, respectively (**Figure 3C** and **3D**). Moreover, the Random Forest model ranked these 17 autoantibodies based on their ability to discriminate between healthy controls and COVID-19 disease severity groups. Follow-up analysis indicated CXCR3-Ab, AT1R-Ab, MASR-Ab, M5R-Ab, and Brady-R1-Ab as the five most significant predictors of COVID-19 classification based on the number of nodes and gini-decrease criteria for measuring variable importance (**Figure 3E** and **3F**). However, other autoantibodies such as PAR1-Ab and STAB-1-Ab were also strong predictors of COVID-19 severity. The interaction between anti-CXCR-3 and anti-AT1R autoantibodies was the most frequent interaction occurring in the decision trees obtained by the Random Forest model (**Supp. Figure 2A and 2B)**. Altogether, these results show that autoantibodies targeting GPCRs and COVID-19 associated molecules perform well as predictors of COVID-19 disease severity and raises the question of whether these autoantibodies against key functional molecules play a significant role in COVID-19 pathophysiology.

### Disruption of autoantibody correlation signatures in severe forms of COVID-19

We have recently reported that hierarchical clustering signatures of anti-GPCR autoantibody correlations are associated with physiological and pathological immune homeostasis^41^. Based on this concept, we investigated the correlation signatures in healthy controls and patients with COVID-19 to explore if changes in autoantibody relationships correlate with disease burden. Bivariate correlation analysis revealed a progressive loss of normal correlation signatures from mild to oxygen-dependent COVID-19 patients. In other words, patients with mild COVID-19 exhibited only minimal differences in the autoantibody correlation signatures when compared to healthy controls (**Figure 4A**). Patients with moderate COVID-19 started to clearly exhibit new relationships among autoantibodies while the severe group displayed the most different topological correlation pattern. Topologically, a positive correlation predominated among the autoantibodies. Of note, autoantibodies targeting nine different molecules presented significant changes in the total correlation distribution, which was determined by the distribution of a pairwise correlation between autoantibodies (**Figure 4B**). In summary, while the autoantibody network signatures were relatively conserved in patients with mild COVID-19 compared to healthy controls, these were disrupted in moderate and most perturbed in severe patients (**Supp. Figure 3)**

**Figure 5:**
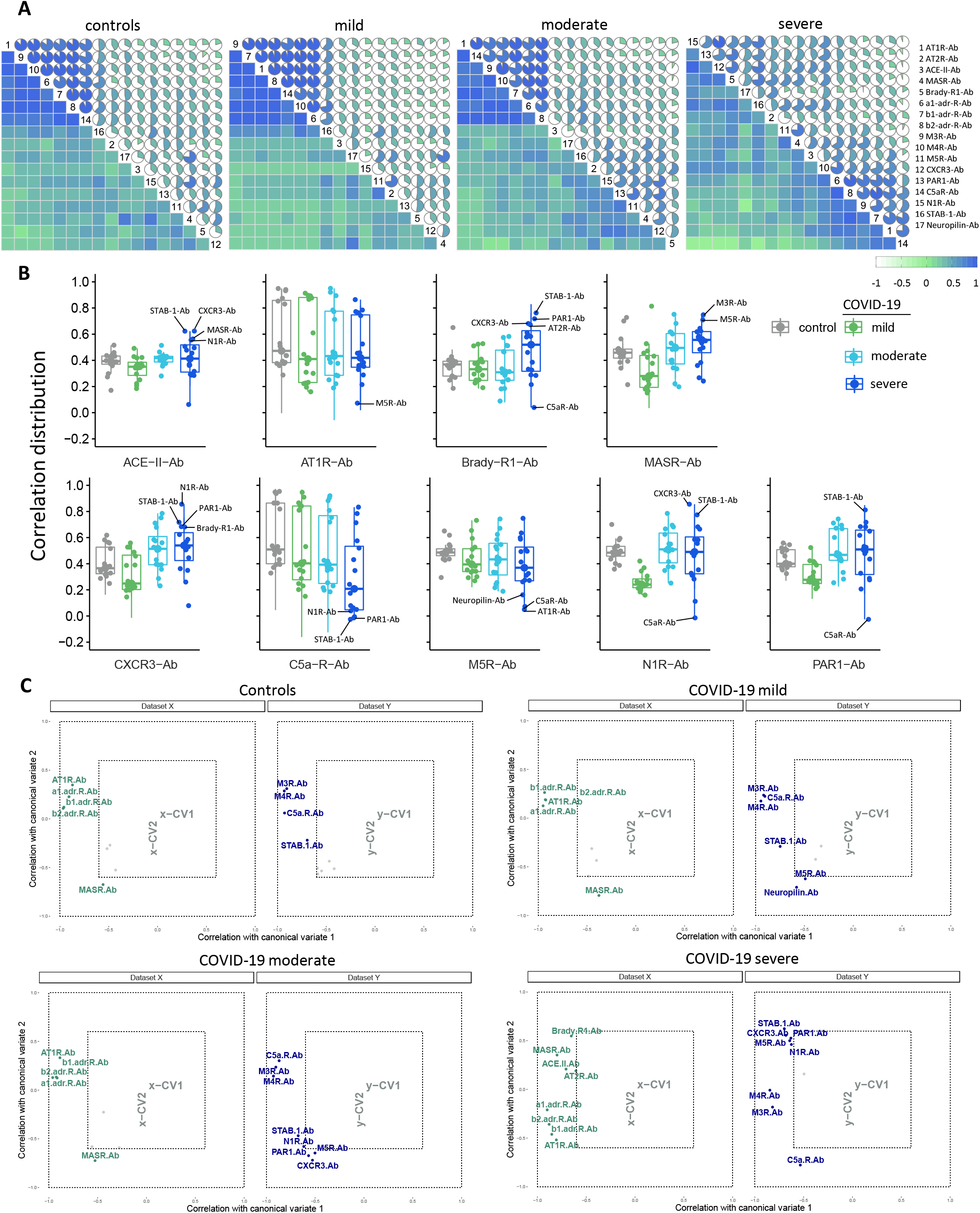
Autoantibody correlation signatures associate with disease burden. (**A**) Correlation matrices of the 17 antibodies (denoted by numbers as per legend) for controls and COVID-19 groups. The color scale bar represents the Spearman’s rank correlation coefficient. (**B**) Box plots of autoantibodies with significantly changes in correlations in relation to the other 16 autoantibodies: those belonging to the RAS are placed in upper row; autoantibodies targeting the other GPCR are exhibited in the lower row. Antibodies with highest or lowest correlations and thus contributing more to changes in the correlation pattern of the severe COVID-19 group are denoted. (**C**) Canonical-correlation analysis (CCA of autoantibodies. Correlation between autoantibodies against molecules belonging or influencing the RAS (dataset X, in green) *versus* the other autoantibodies (those targeting th other GPCRs, neuropilin, and STAB1; dataset Y, in blue). Autoantibodies corrrelations are plotted based on their corrrelation (that range from −1 to 1) with the first 2 canonical variates (x-CV1 and xCV2 or y-CV1 and y-CV2). Note, that this values ranging from −1 to 1 are not the same as the Spearman’s rank correlation coefficient. Only autoantibodies with a correlation **≥** 0.6 of Spearman’s rank correlation coefficient are shown while those with a correlation < 0.6 (grey points) have their names omitted.

To better understand these changes in autoantibody correlation signatures, we performed canonical-correlation analysis (CCA), which is a multivariate statistical method to determine the linear relationship between two groups of variables^42^. CCA was carried out splitting autoantibodies into two groups: those against molecules belonging or influencing the RAS (Dataset X) compared with those autoantibodies targeting other GPCRs, neuropilin, and STAB1 (Dataset Y). This approach confirmed the changes in autoantibody relationship patterns revealed by Bivariate correlation analysis. In addition, the CCA indicated changes based on COVID-19 severity are in agreement with the Random Forest model. For instance, in this multivariate correlation approach autoantibodies targeting CXCR3 showed Spearman’s rank correlation coefficient >0.6 only in the moderate and severe groups (**Figure 4C**). In this context, while Brady-Ab only appeared in the severe group, AT1R-Ab, MASR-Ab, and M5R-Ab exhibited changes in their correlation patterns that were only observed in the severe group.

## DISCUSSION

The precise mechanisms by which the SARS-CoV-2 infection triggers the production of autoantibodies remains unknown. However, a potential antigenic cross-reactivity (molecular mimicry) between SARS-CoV-2 and human tissues has been hypothesized^43–48^. Furthermore, the hyperinflammatory reaction triggered by this virus results in tissue damage causing systemic immune-related manifestations that have been reported in patients with COVID-19^49^. When compared with patients manifesting mild disease, those with moderate COVID-19 symptoms, present with strong antibody production and high titers of neutralizing antibody^50^, but, as shown here, also with an increased production of autoantibodies. Thus, our work reinforces the concept that SARS-CoV-2 infection may trigger a life-threatening autoimmune disease, suggesting that this occurs against multiple functional molecules with key functions in immune and vascular homeostasis^1–3,51^. While we found no differences in the levels of autoantibodies against neuronal receptors such as ADRA1A, ADRB1, ADRB2, CHRM3, and CHRM4, those targeting other GPCRs and RAS-related molecules were significantly dysregulated when comparing controls with moderate and severely affected COVID-19 patients. In this context, while we have previously reported that anti-AT1R has agonist properties^13,41,52,53^, the mechanistic action of the other autoantibodies we identified remains to be investigated. For instance, we hypothesize that antibodies against CXCR3 might block the migration of immune cells that express CXCR3, such as natural killer cells, as well as CD4+ and CD8+ T cells that are critical for the killing of viruses in the lung^54–57^. However, if antibodies against CXCR3 have agonistic properties, potentializing CXCL9/CXCL10/CXCL11 signaling, they could exacerbate deleterious hyperinflammation. Anyhow, the results of our work underscores recent studies^3,6,10,12^ that report the generation of autoantibodies following SARS-CoV-2 infection. Importantly, our data indicate that an additional immunological layer is present where autoantibodies targeting GPCRs and RAS-related molecules are associated with COVID-19 burden. This association sheds new light on the proposed immuno-hematological mechanisms underlying the development of COVID-19 infection, which is based on the abnormal activation of the ACE-II/Angiotensin II (Ang II)/AT1R/RAS axis together with a reduction of the ACE-II/ Angiotensin-(1-7)/MASR branch occurring together with several immunological dysregulation events^58^.

The Random Forest model revealed the interaction between autoantibodies targeting CXCR3 and AT1R as the most important predictors of COVID-19 severity. There is an essential biological connection between CXCR3 and AT1R. Blocking AT1R impairs the release of several chemokines, including CXCL10, the ligand for CXCR3^59^, a chemokine receptor highly expressed by effector T cells controlling the traffic and function of CD4+ and CD8+ T cells during inflammation^60–63^. Further, CXCR3 has been strongly associated with both autoimmune and inflammatory diseases^64^. Meanwhile, increased levels of Ang II together with the hyperactivation of its receptor (AT1R) have been associated with unfavorable COVID-19 disease^16,65^. This pathological mechanism has been explored as a therapeutic approach for COVID-19 by clinical trials with Losartan, an AT1R antagonist^8,66^. AT1R orchestrates several important immunological functions and Losartan treatment has been previously demonstrated to have immunomodulatory properties. Ang II is the main effector molecule of RAS that upon binding to AT1R promotes vasoconstriction, inflammation, oxidative stress, coagulation, and fibrosis, all playing an important pathological role during SARS-CoV-2 infection^19^.

Furthermore, our work indicates a change in the relationship between autoantibodies targeting GPCRs and RAS that associate with COVID-19 severity, which was shown by increasing disruption of autoantibody correlations according disease burden. This observation provides new insights into the biology of autoantibodies, which is in line with our previous observation that GPCR-specific autoantibody signatures associate with physiologic and pathologic immune homeostasis^41^. Further, as several epitopes on highly interrelated GPCRs are likely overlapping^67^, a change in the correlation structure might indicate that new epitopes are targeted in severe COVID-19. These epitopes could have different functional properties. However, this also represents a limitation of our work that demand future investigations. Although we have previously assessed how these autoantibodies act in the context of systemic autoimmune diseases^13,41,52,68–72^, mechanistic investigations are missing to characterize how all these autoantibodies can simultaneously affect (i.e., stimulating or blocking) their targets in the context of COVID-19. For instance, future evaluation will be necessary to determine if they have synergistic effects in the presence of endogenous ligands as maybe the case with anti-CXCR3 autoantibodies and CXCL9/CXCL10/CXCL11. Since GPCRs comprise the largest superfamily of integral membrane proteins in humans^14^, it is also possibly that several additional anti-GPCR autoantibodies remain to be discovered. Likewise, several SARS-CoV-2 strains have been identified^73^ and it will be important to investigate whether they induce different autoantibody patterns that may contribute to disease outcomes. Of note, autoantibodies are present in healthy individuals and immunization with GPCR-overexpressing membranes can induce the production of autoantibodies targeting GPCRs^41^. Thus, another important issue to be addressed is whether the recently developed vaccines against COVID-19^74^ could induce the production of anti-GPCR autoantibodies.

In conclusion, this study identifies new autoantibodies which are dysregulated by SARS-CoV-2. Our data also indicates that anti-GPCR antibodies represent potential new clinically relevant biomarkers that predict COVID-19 severity. The disruption of autoantibody network signatures in severe patients suggests a progressive loss of autoantibody homeostasis that accompanies the progression of the disease triggered by SARS-CoV-2-induced immune dysregulation. Since a better understanding of the COVID-19 pathogenesis may open new avenues to improve diagnostic options^75,76^, the results reported here may provide new insights to improve the clinical management of COVID-19 patients.

## ONLINE METHODS

### Patient Cohort

We included 246 adults from Jewish communities across 5 states of the United States of America, who had developed symptomatic COVID-19 disease before receiving any SARS-CoV-2 vaccine and participated in an online survey developed to determine the most common symptoms and outcomes of SARS-CoV-2 infection^77,78^. Details about the survey study, patient demographics and symptoms have been previously described^77,78^. 77 randomly selected age- and sex-matched healthy controls (SARS-CoV-2 negative and without symptoms of COVID-19) were included in this study and their autoantibody data were compared to 169 individuals who were SARS-CoV-2 positive (determined by positive nasopharyngeal swab). The SARS-CoV-2 infected cohort formed the COVID-19 mild (n=74; fever duration ≤ 1 day; peak fever of 37.8 C), COVID-19 moderate (n=63; fever duration ≥ 7 day; peak fever of ≥ 38.8 C) and COVID-19 severe (n=32; severe symptoms and requiring supplemental oxygen therapy) groups. Disease severity for SARS-CoV-2 positive individuals was based on the World Health Organization (WHO) severity classification. All healthy controls and patients provided written consent to participate in the study, which was performed in accordance with the Declaration of Helsinki and approved by the IntegReview institutional review board. In addition, this study followed the Strengthening the Reporting of Observational Studies in Epidemiology (STROBE) reporting guideline.

### Detection of IgG autoantibodies

Human IgG autoantibodies against 14 different GPCRs (AT1R, AT2R, MASR, Brady-R1, alpha1-adr-R, beta1-adr-R, beta2-adr-R, M3R, M4R, M5R, CXCR3, PAR1, C5a-R, N1R), 2 molecules serving as entry for SARS-CoV-2 (ACE-II, neuropilin), as well as antibodies against the transmembrane receptor STAB-1 were detected from frozen serum using commercial ELISA Kits (CellTrend, Germany) according to manufacturer’s instructions, as previously described^79^. Briefly, duplicate samples of a 1:100 serum dilution were incubated at 4 °C for 2 h. The autoantibody concentrations were calculated as arbitrary units (U) by extrapolation from a standard curve of five standards ranging from 2.5 to 40□U/ml. The ELISAs were validated according to the Food and Drug Administration’s Guidance for Industry: Bioanalytical Method Validation.

### Interaction network and enrichment analysis of autoantibody targets

The interaction network of 17 GPCR-autoantibody targets was built using the online tool string^80^ (https://string-db.org/). Gene ontology (GO) enrichment analysis of the 17 autoantibody targets was performed using GO Biological Process 2021 analysis through the Enrichr webtool^81–83^. Circos Plot of antibody targets and pathway association was built using Circos online tool^84^.

### Differences in autoantibody levels

Box plots showing the different expression levels of 17 anti-GPCR-autoantibodies from COVID-19 patients (mild, severe and oxygen-dependent groups) and healthy controls were generated using the R version 4.0.5 (The R Project for Statistical Computing. https://www.r-project.org/), R studio Version 1.4.1106 (R-Studio. https://www.rstudio.com/), and the R packages ggpubr, lemon, and ggplot2. Statistical differences of autoantibody levels were assessed using t-test.

### Principal Component Analysis

Principal Component Analysis (PCA) with spectral decomposition^38,39^ was used to measure the stratification power of the 17 autoantibodies to distinguish between COVID-19 (mild, moderate and severe patients) and healthy controls. PCA was performed using the R functions prcomp and princomp, through factoextra package (Principal Component Analysis in R: prcomp vs princomp. http://www.sthda.com/english/articles/31-principal-component-methods-in-r-practical-guide/118-principal-component-analysis-in-r-prcomp-vs-princomp/).

### Machine learning model and autoantibody ranking

We employed Random Forest model to construct a classifier able to discriminate between controls, mild, severe, and oxygen-dependent COVID-19 patients. This approach aimed to identify the most significant predictors for severe COVID-19. We trained a Random Forest model using the functionalities of the R package randomForest (version 4.6.14)^85^. Five thousand trees were used, and the number of variables resampled were equal to three. Follow-up analysis used the Gini decrease, number of nodes, and mean minimum depth as criteria to determine variable importance. The adequacy of the Random Forest model as a classifier was assessed through out of bags error rate and ROC curve. For cross-validation, we split the dataset in training and testing samples, using 75% of the observations for training and 25% for testing.

### Autoantibody correlation signatures: Bivariate and multivariate correlation analysis

Bivariate correlation analysis of autoantibodies for each group (controls, mild, severe, oxygen-dependent COVID-19 patients) was performed using the corrgram, psych, and inlmisc R packages. In addition, multilinear regression analysis of relationships between different variables (auto-antibodies) was performed using the R packages ggpubr, ggplot2 and ggExtra. Circle plots were also build using the R packages qgraph, ggplot2, psych, inlmisc to visualize the patterns of Spearman’s rank correlation coefficients between autoantibodies. CCA^86^ of autoantibodies against molecules associated with RAS, other GPCRs and SARS-CoV-2 entry molecules was performed using the R packages CCA and whitening^86^. CCA is a classic statistical tool to perform multivariate correlation analysis. We used log-transformed antibody levels to carry out both bivariate correlation and CCA analysis.

## Data Availability

A reporting summary for this article is available as a Supplementary Information file. The source data underlying the Main and Supplementary Figures are provided as a Source Data file. All R packages used in this manuscript are described in the Reporting Summary.

## Acknowledgments

We acknowledge the patients for the participation in this study. We also thank the São Paulo Research Foundation (FAPESP grants 2018/18886-9, 2020/01688-0, and 2020/07069-0 to OCM; 2020/09146-1 to PPF) for financial support. Computational analysis was supported by FAPESP. This work was supported by the Deutsche Forschungsgemeinschaft (DFG) founding the excellence cluster Precision Medicine in Inflammation, project TI4 and CD1, by the COVID fond of Schleswig-Holstein as well as the DFG project RI 1056 11-1/2.

## Contributions

OCM, LFS, GH, AZR, GR, YS wrote the manuscript; OCM, LFS, AZR, SS, JYH, AM, TL, LMG, GH, HG, AS, AMH, HDO, GR, YS provided scientific insights; COM, LFS, YBL, AHC, ISF, DRP, GCB, PPF, DLMF performed data and bioinformatics analyses; JIS, AZR, IZ, MTL diagnosed, recruited or followed-up the patients; HH, JJ, CM, KSF coordinated the serum collection, databank or performed the experiments. HH, AH, HA, JZ, AZR, GR and YS conceived the project and designed the study; OCM, GH, LFS, AZR, TL, LMG, YBL, GR, YS, HDO revised and edited the manuscript; GR and YS supervised the project.

## Competing interests

The authors declare that H.H. and K.S.F. are CellTrend managing directors and that GR is an advisor of the company CellTrend and earned an honorarium for her advice between 2011 and 2015. The other authors declare no competing interests.

---

**Supp. Fig 1.**
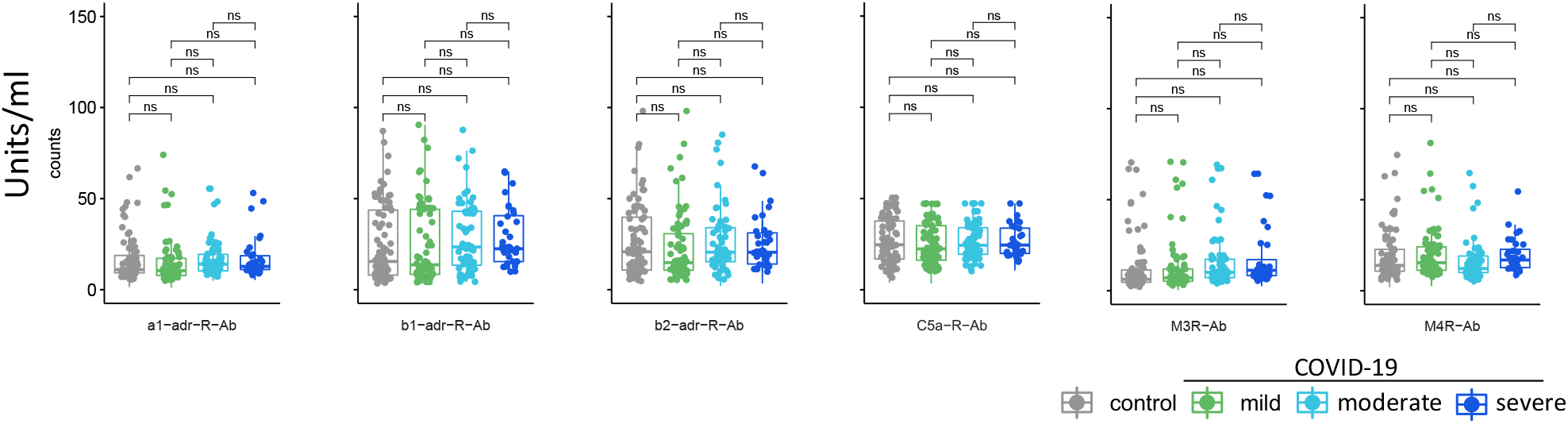
Autoantibodies against GPCRs which do not differentiate between COVID-19 and healthy controls. Box plots of 6 antibodies which were not significantly different between COVID-19 patient groups and healthy controls.

**Supp. Fig 2.**
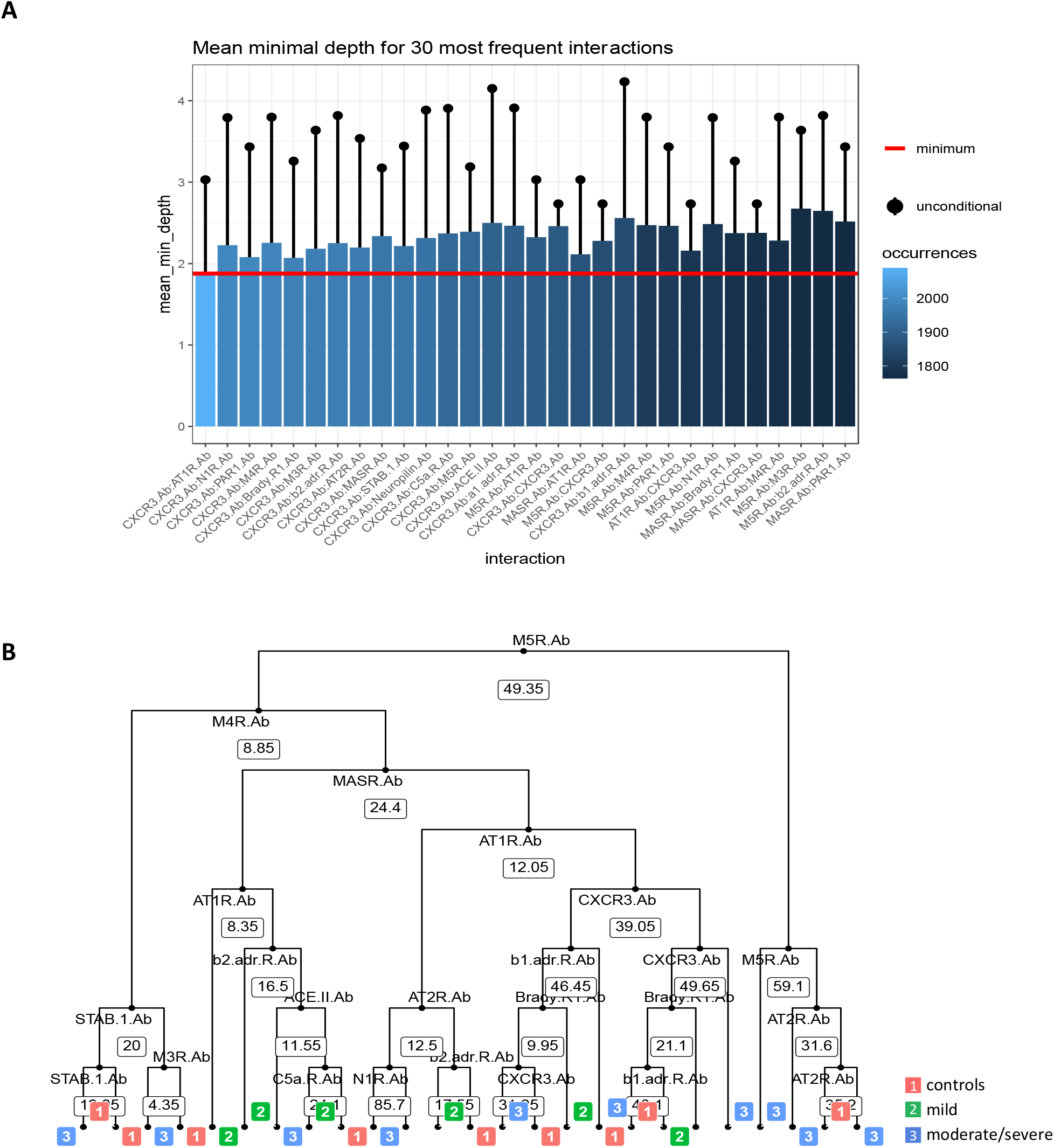
Random Forest shows interactions between variables and decision tree. (**A**) interactions are ordered by decreasing number of occurrences. Thus, the plot shows from the most frequent occurring interactions between the autoantibodies on the left side with lighter blue color to the least frequent occurring interactions on the right side of the graph with darker blue color. The horizontal red line indicates the mean minimum depth and the black lollipop indicate the unconditional mean minimal depth of a variable. (**B**) Classification tree obtained by random forest model as described in **Figure 3**. This is the tree decision with the least number of nodes from the 5000 trees obtained by Random Forest analysis.

**Supp. Fig 3.**
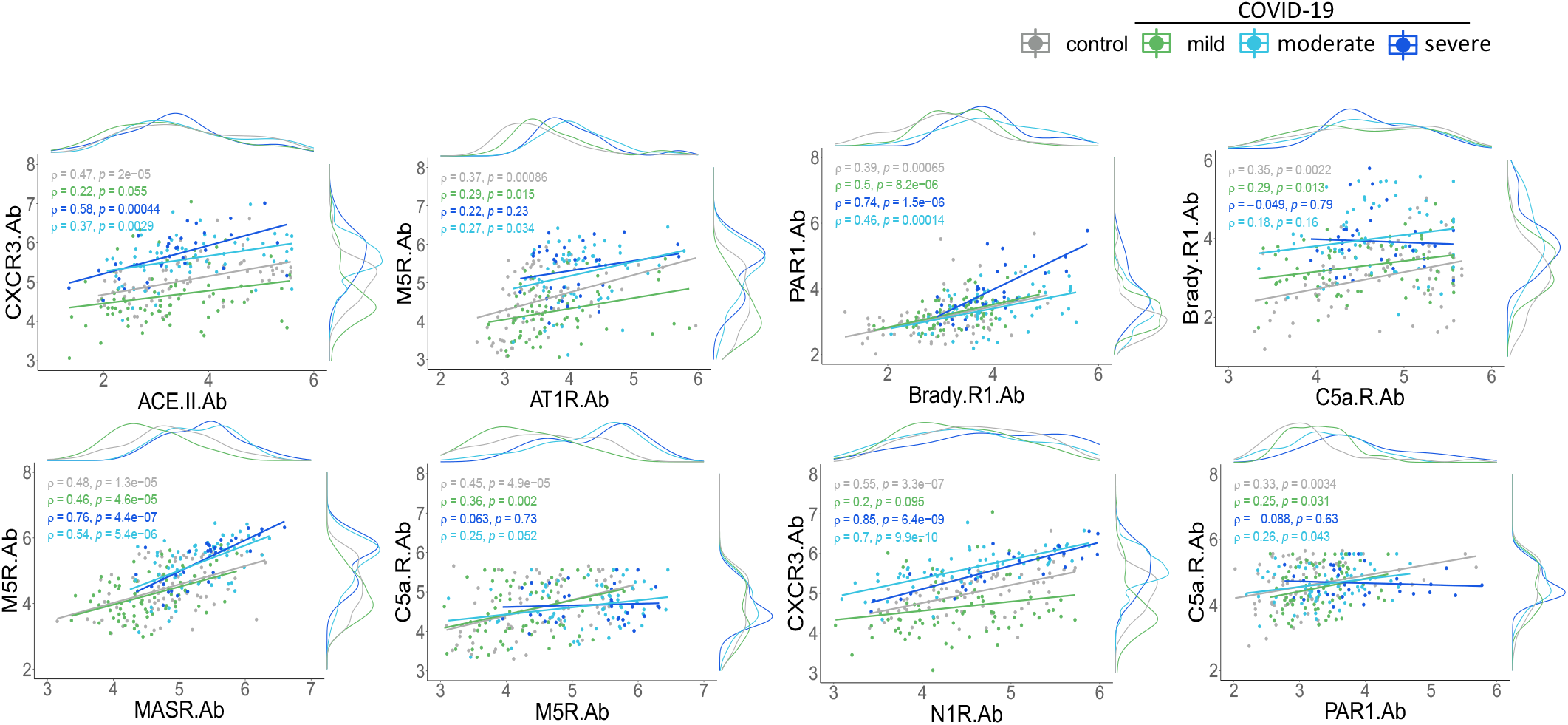
Scatter plots with marginal density plots display the relationship between different autoantibodies. Correlation coefficient (ρ) and significance level (p-value) for each correlation are shown within each graph.

**Supp. Fig 4.**
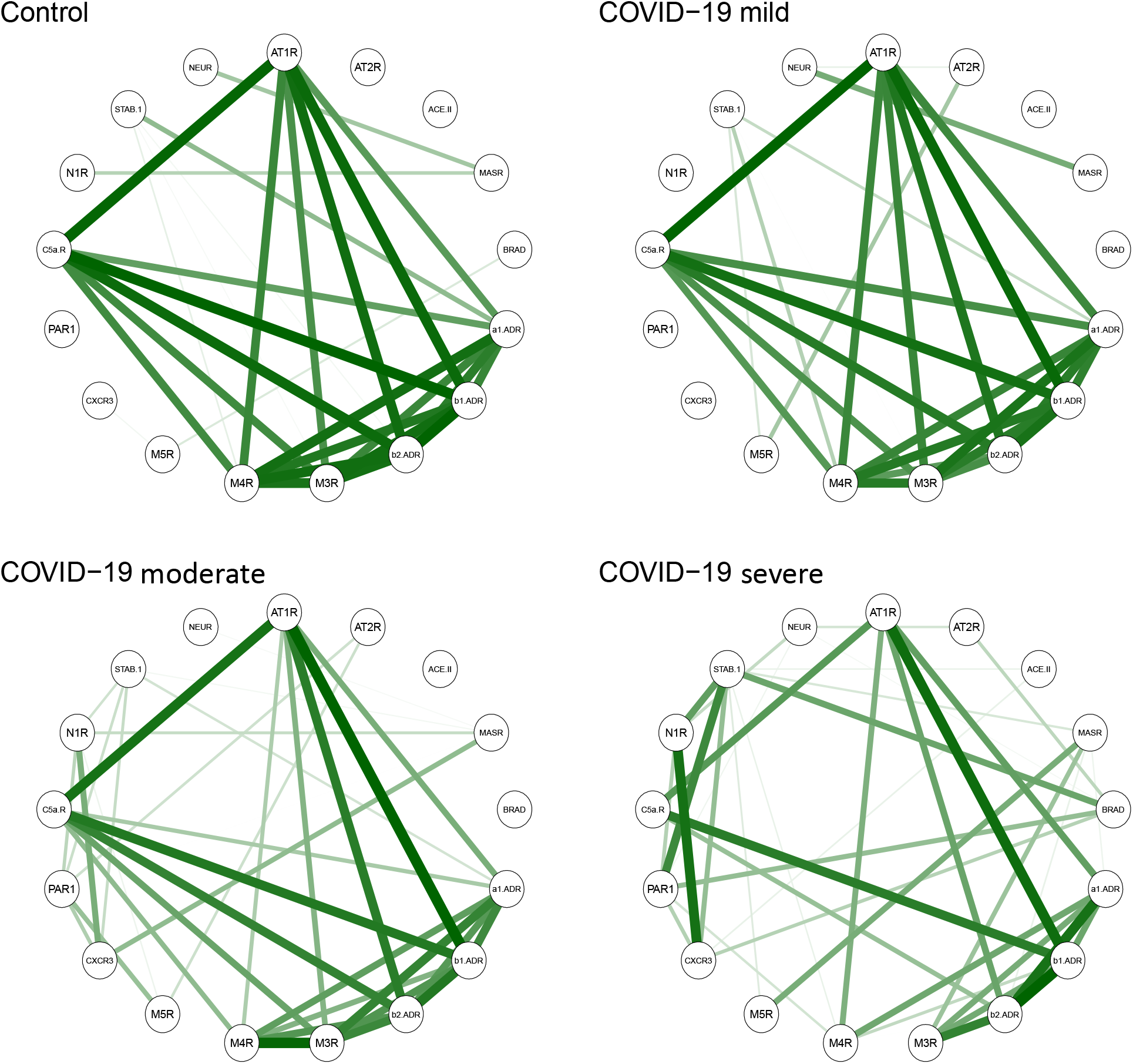
Effects of COVID-19 severity on autoantibody correlations. The relationships among the different autoantibodies in sera from healthy controls, COVID-19 mild, moderate, and severe patient groups as shown by circular networks based on Spearman’s rank correlation coefficients for autoantibodies. The nodes in the graphs represent variables (each autoantibody), and a line between two nodes indicates the Spearman’s rank correlation coefficient. The line width indicates the strength of the association, with stronger correlations indicated by thicker and greener lines. Only correlations >0.6 are shown. Multiple connections of nodes indicate clustering.

## Notes

### Author Declarations

All healthy controls and patients provided written consent to participate in the study, which was performed in accordance with the Declaration of Helsinki and approved by the IntegReview institutional review board. In addition, this study followed the Strengthening the Reporting of Observational Studies in Epidemiology (STROBE) reporting guideline.

